# How to enlarge lesion size in temperature control mode using an irrigation catheter with a flexible-tip of laser-cut kerfs

**DOI:** 10.1101/2023.08.08.23293854

**Authors:** Masateru Takigawa, Junji Yamaguchi, Masahiko Goya, Hidehiro Iwakawa, Tasuku Yamamoto, Hidehiro Iwakawa, Miki Amemiya, Takashi Ikenouchi, Miho Negishi, Iwanari Kawamura, Kentaro Goto, Takatoshi Shigeta, Takuro Nishimura, Tomomasa Takamiya, Susumu Tao, Katsuhiro Ohuchi, Sayaka Suzuki, Shinsuke Miyazaki, Tetsuo Sasano

## Abstract

**Background & Objectives:** The TactiFlex^TM^ SE irrigation catheter installs a laser-cut flexible tip with a thermocouple embedded 0.3mm from the tip, being used with temperature-control mode. We aimed to elucidate a safer method to enlarge the lesion size using this catheter.

**Methods:** RF-applications were performed at a range of powers (35W, 40W, and 45W), CFs (10g/20g), and durations (60s, 120s, and 180s) using perpendicular/parallel catheter orientation with normal saline irrigation (NS-irrigation) and Half NS-irrigation (HNS-irrigation) in an ex-vivo model (Step 1). Further, similar RF-applications of 35W/40W/45W for 60s/120s/180 in NS-irrigation and 35W/40W for 60s/120s/180s in HNS-irrigation) were performed in 4 swine (Step 2). Lesion characteristics and the incidence of steam-pops were assessed.

**Results:** In this ex-vivo experiment, 47(16.3%) steam-pops (13 in NS-irrigation vs 34 in HNS-irrigation, p=0.001) were observed in 288 lesions. In NS-irrigation group, the most aggressive setting (45W/180s, n=7/13, 54%) played a large part of steam-pop occurrence. Lesion size proportionally increased with longer ablation duration, but not with HNS-irrigation. The optimal cut-off of %impedance-drop to predict steam-pops was 20% with the sensitivity, specificity, positive predictive value (PPV), and negative predictive value (NPV) of 78.3%, 80.6%, 43.4%, and 95.1%, respectively. In the in-vivo experiment, steam-pops were not observed with NS-irrigation (n=0/35), but were with HNS-irrigation (6/21, 29%). Lesion size proportionally increased with longer ablation duration, but not with HNS-irrigation partially because of the higher incidence of temperature-guard use (p=0.07). The optimal cut-off of %impedance-drop to predict steam-pops was 24% with the sensitivity= 83.0%, specificity=52.0%, PPV= 17.2%, and NPV= 96.3%, respectively, but 20% may be used with a safety margin.

**Conclusions:** Although this novel catheter is generally safe, increasing ablation duration instead of using HNS-irrigation may be recommended for larger lesions.

## Introduction

Radiofrequency (RF) catheter ablation is an established treatment for cardiac arrhythmias. Optimal lesion formation and avoidance of excessive heating are necessary for effective ablation^1^. However, lesions produced by using standard RF-ablation techniques can be limited in depth, thus challenging efficacy of ablation when the region of interest is contained deep within myocardium^2^, such as interventricular septum, left ventricular summit, midmyocardial sites, and papillary muscles^3^. Prolonged, high-power RF-ablation, irrigation of half-normal saline, coronary arterial or venous ethanol injection, RF-ablation via a needle catheter, and bipolar ablation, may be optional strategies.^4–5^ However, these techniques require special expertise and equipment that are not widely available, and their feasibility and efficacy may be limited.

Multiple factors including power, duration, catheter-tissue contact force (CF), electrode diameter, circuit impedance, irrigation flow, tissue thickness, and myocardial blood flow have been reported to be associated with lesion characteristics^6,7^. The TactiFlex^TM^ SE is a novel irrigation catheter installed with a laser-cut flexible tip with a CF sensor. Although the catheter embeds only a single thermocouple, but within the extreme proximity from the tip (0.3mm from the tip) for accurate temperature monitoring, and effective convective cooling is achieved by efficient irrigation flow irrespective of the tissue-catheter interface angle, which allows a safer and more continuous RF-application with temperature control setting. We aimed to elucidate the strategy to safely create deep lesions using this novel technology.

## Methods

### RF applications

A calibrated roller pump (CoolPoint, Abbott Medical) connected to the catheter delivered saline solution at 13 ml/min during RF delivery, using the TactiFlex™ SE contact-sensor ablation catheter which has a laser-cut 4- mm flexible 8Fr tip, irrigated from the proximal to distal end through laser-cut kerfs, and from four holes on the distal end of the tip. An Ampere RF generator (St. Jude Medical) was connected to deliver 550 kHz unmodulated sine-wave RF energy pulses in a temperature-controlled mode (maximum temperature 43℃). Different power settings, contact intensities, RF duration, and catheter orientation were used during the experiment, as discussed in the following sections.

### Ex-vivo experiment with Swine excised hearts

Commercially obtained swine hearts (N = 48 in total) excised within <24h were preserved in a fresh state and used for this ex-vivo experimental model. A section of the porcine left ventricular myocardium was placed on the ground plate in the circulating saline bath with 5.0 L saline at 37°C. (Figure 1A). To simulate the clinical setting, 0.2 m/s flow pump was used for simulating the left atrial inflow velocity, and the salinity was controlled to maintain the impedance level of 100+/-5ohm, measured by the catheter above the myocardial slab.

**Figure 1.**
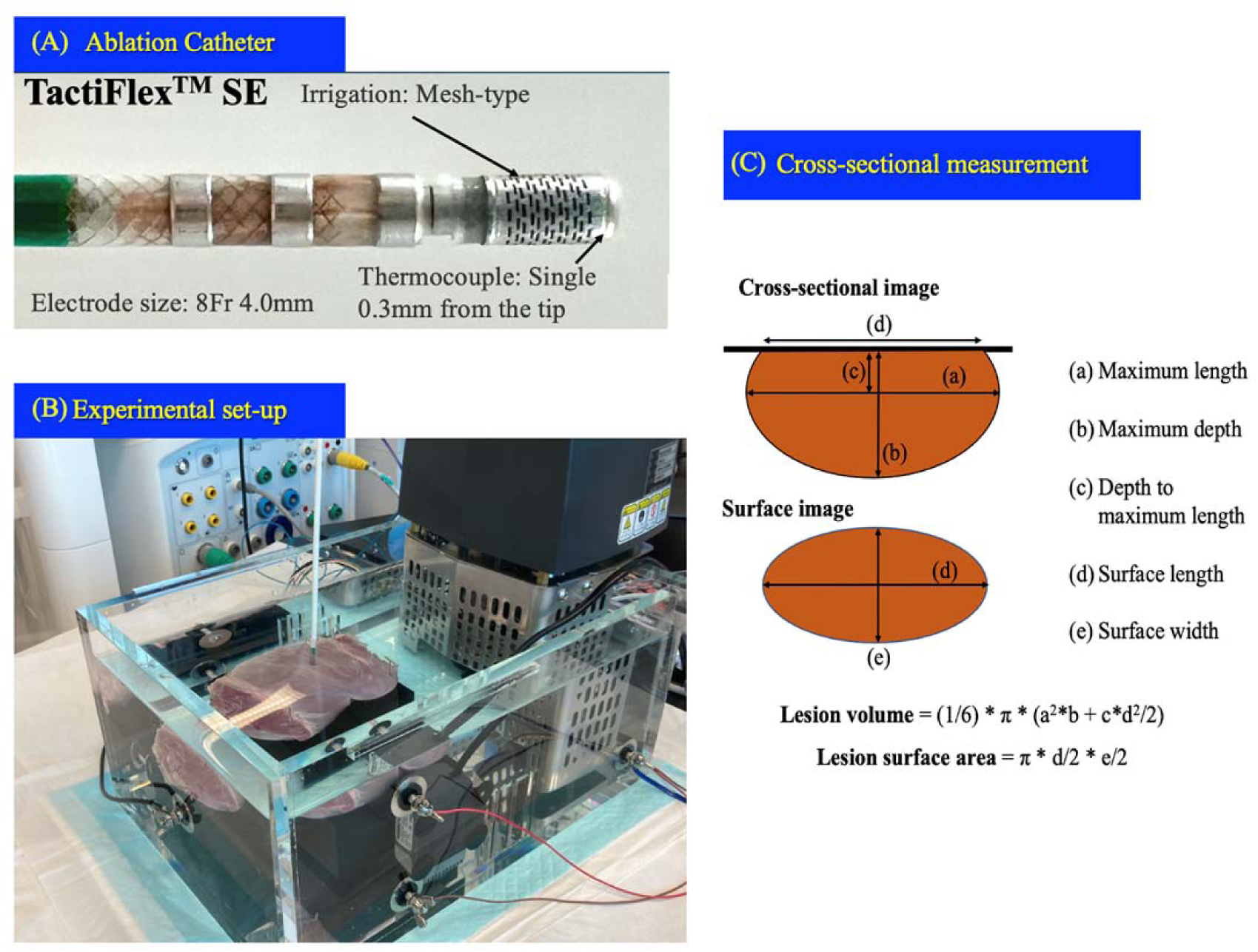
(A) Catheter design of the TactiFlex^TM^ SE ablation catheter with contact sensor, installing a laser-cut 4-mm flexible 8Fr tip, which is irrigated from the proximal to distal end through the laser-cut kerfs. A single thermocouple embedded within 0.3mm from the tip for temperature monitoring, enabling ablation with temperature control mode. (B) Ex-vivo experimental model. (C) Scheme of the surface and cross-sectioned lesion.

To obtain a range of lesion sizes, myocardial lesions were created at separate sites with CF set at 10g and 20g, and power settings set at 35, 40, and 45W. For each CF and power setting, RF delivery was delivered for 60s, 120s and 180 seconds. The catheter was placed perpendicular and parallel to the tissue with different irrigations including normal saline (NS) and half normal saline (HNS) (Figure 1B). Four lesions for each combination of settings were created (288 lesions in total).

### In-vivo experiment with Swine

The protocol of the in-vivo experiment was approved by the Institutional Animal Care and Use Committees of Tokyo Medical and Dental University (A2022-179A). Four Swine (2 female, age 3-4 months; weight 50-65 kg) were sedated with an intramuscular injection of ketamine hydrochloride (10 mg/kg) and xylazine(2mg/kg). After an inhalation of isoflurane, each Swine was intubated, and anesthesia was maintained with 2%–3% isoflurane. Swine were ventilated with a respirator (Carestation/Carescape, GE Health-care, Chicago, IL), using room air supplemented with oxygen. An intravenous sheath was inserted at the internal jugular vein for drug and fluid infusion. Amiodarone was dripped intravenously to prevent ventricular arrhythmias. Arterial blood gases were monitored periodically, and ventilator parameters were adjusted to maintain blood gases within physiological ranges.

A decapolar catheter was placed in the coronary sinus (CS) through the internal jugular vein. Right and left ventricular mapping with the Advisor™ HD Grid Mapping Catheter, Sensor Enabled™ (Abbott) was performed during CS pacing, followed by radiofrequency applications using TactiFlex^TM^ SE. Power setting ranges of 35, 40, and 45W, and ablation duration of 60s, 120s and 180 seconds, with CF of 10-20g were scheduled with NS-irrigation. The same protocol was scheduled with HNS-irrigation, but only with 35W and 40W applications. In total, 60 lesions were planned.

### Lesion assessment

After RF delivery, lesion surface was measured as shown in Figure 1C. The myocardium was cross-sectioned along the surface length at the level of each lesion. The cross-sectioned area was also measured as shown in Figure 1C. Each lesion was measured with a digital caliper with a resolution of 0.1 mm by one observer; the observer was blinded to the lesion protocol. Surface area and lesion volume were calculated from the following formulae^8,9,10^:

Lesion volume = (1/6) × π × (e^2^ × d + c× a^2^/2)

Lesion surface area = π × a/2 × b/2

Lesions with steam-pops were excluded from the lesion size analysis.

### Statistical analysis

The data are presented as median [25%ile-75%ile]. Continuous variables were compared by Wilcoxon test or Kruskal-Willis test. Categorical variables were compared by an χ^2^ test or Fisher’s exact test. Significant differences were further evaluated by using Steel-Dwass test for pairwise multiple comparisons. A P-value < 0.05 was considered statistically significant.

## Results

### Incidence of steam-pops and lesion metrics: HNS vs NS

In total, 288 lesions including 47 (16.3%) steam-pops were observed. Generally, steam-pops more frequently occurred while HNS was used for irrigation (13(9.3%) in NS group vs 34(23.6%) in HNS, P=0.001).

Although the incidence of steam-pops tended to increase as the ablation power and ablation duration increased as shown in Figure 2, in the NS-irrigation group, the most aggressive setting (45W/180s, n=7/13, 54%) played a large part of steam-pop occurrence, and the incidence of steam-pops was generally low in the other settings. To avoid the bias due to steam-pop lesions while comparing lesion size between the HNS-group and NS-group, ablation settings causing steam-pops in one cohort were also excluded from the other. In total, 105 lesions in each group were compared, showing that the length, depth, volume, and surface area of the lesion did not differ, although impedance-drop was significantly larger in the HNS group as shown in Figure 3A and 3B.

**Figure 2.**
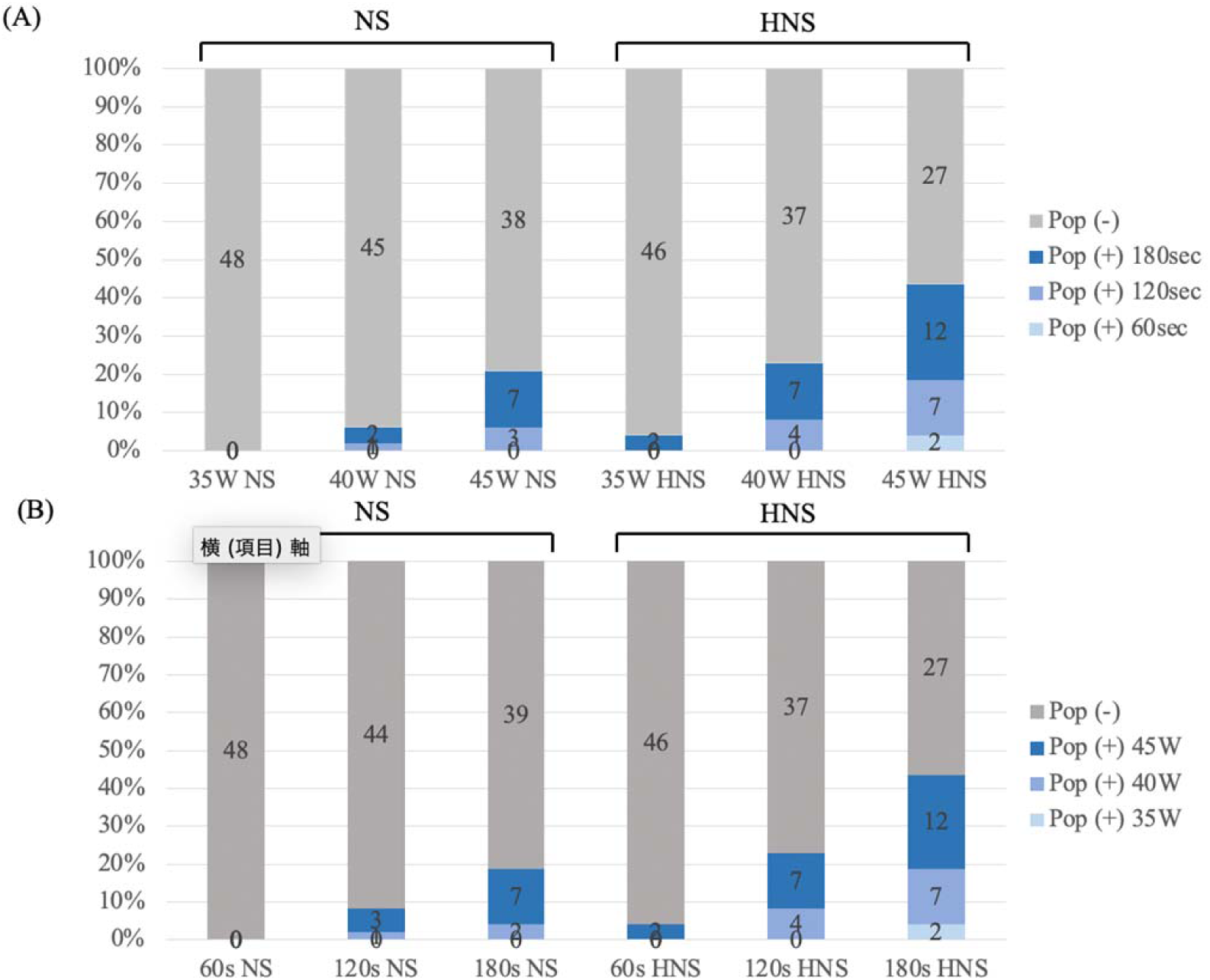
Incidence of steam-pops in ex-vivo condition. **A. Ablation power vs steam-pops** **B. Ablation duration vs steam-pops**

**Figure 3.**
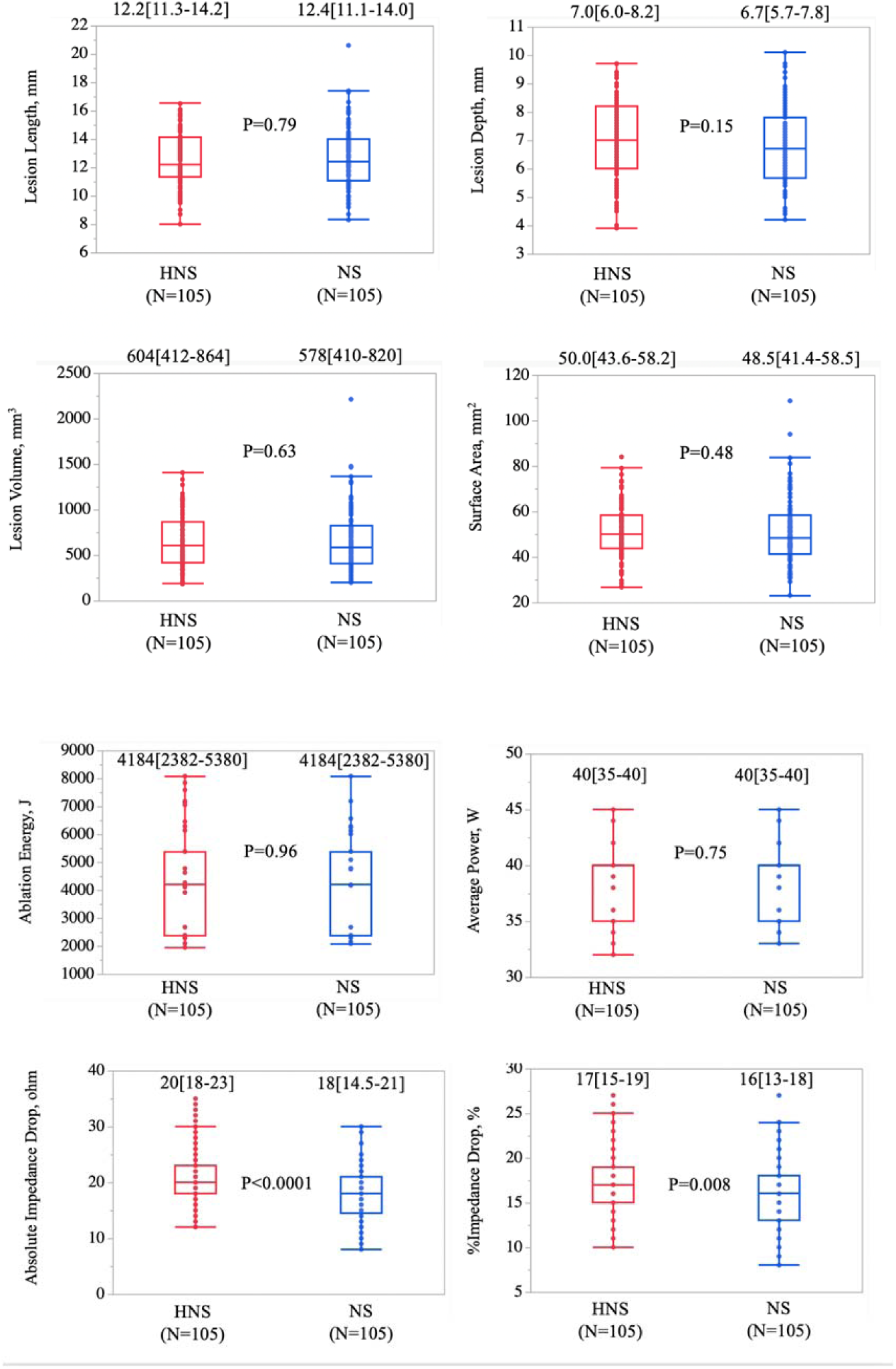
Comparison of lesion metrics and ablation parameters between HNS-irrigation and NS-irrigation in ex-vivo model. **(A) HNS-irrigation vs NS-irrigation: Lesion metrics** **(B) HNS-irrigation vs NS-irrigation: Ablation parameters**

### Incidence of steam-pops and lesion metrics: 60sec vs 120sec vs 180sec

As shown in Figure 2, the incidence of steam-pops increased as the ablation duration and power increased in both the NS-irrigation group and the HNS-irrigation group. Lesion metrics and ablation parameters during RF-applications depending on the ablation duration and power with or without steam-pops are demonstrated in Table 1. Generally, lesion size increased as the duration and power increased. However, to precisely analyze the impact of long duration RF applications, RF-applications with HNS-irrigation was removed, to minimize the bias caused by high-incidence of steam-pops for the comparison of lesion metrics among 60sec, 120sec, and 180sec. In addition, ablation settings causing steam-pops in one cohort were also excluded from the other two cohorts. Finally, 39 lesions in each group were compared, showing that the length, depth, volume, and surface area of the lesion significantly increased as the ablation duration increased as shown in Figure 4A and 4B.

**Figure 4.**
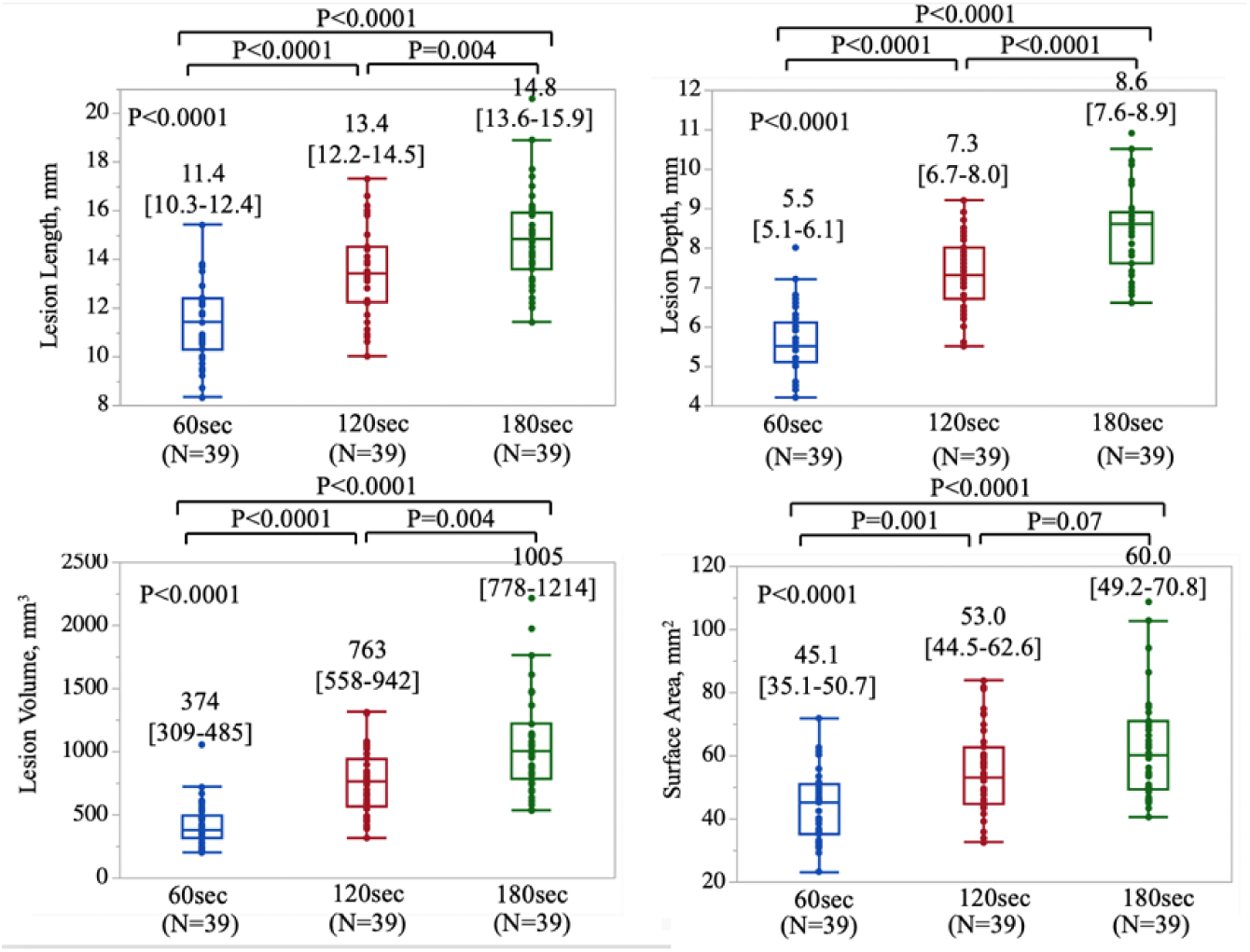
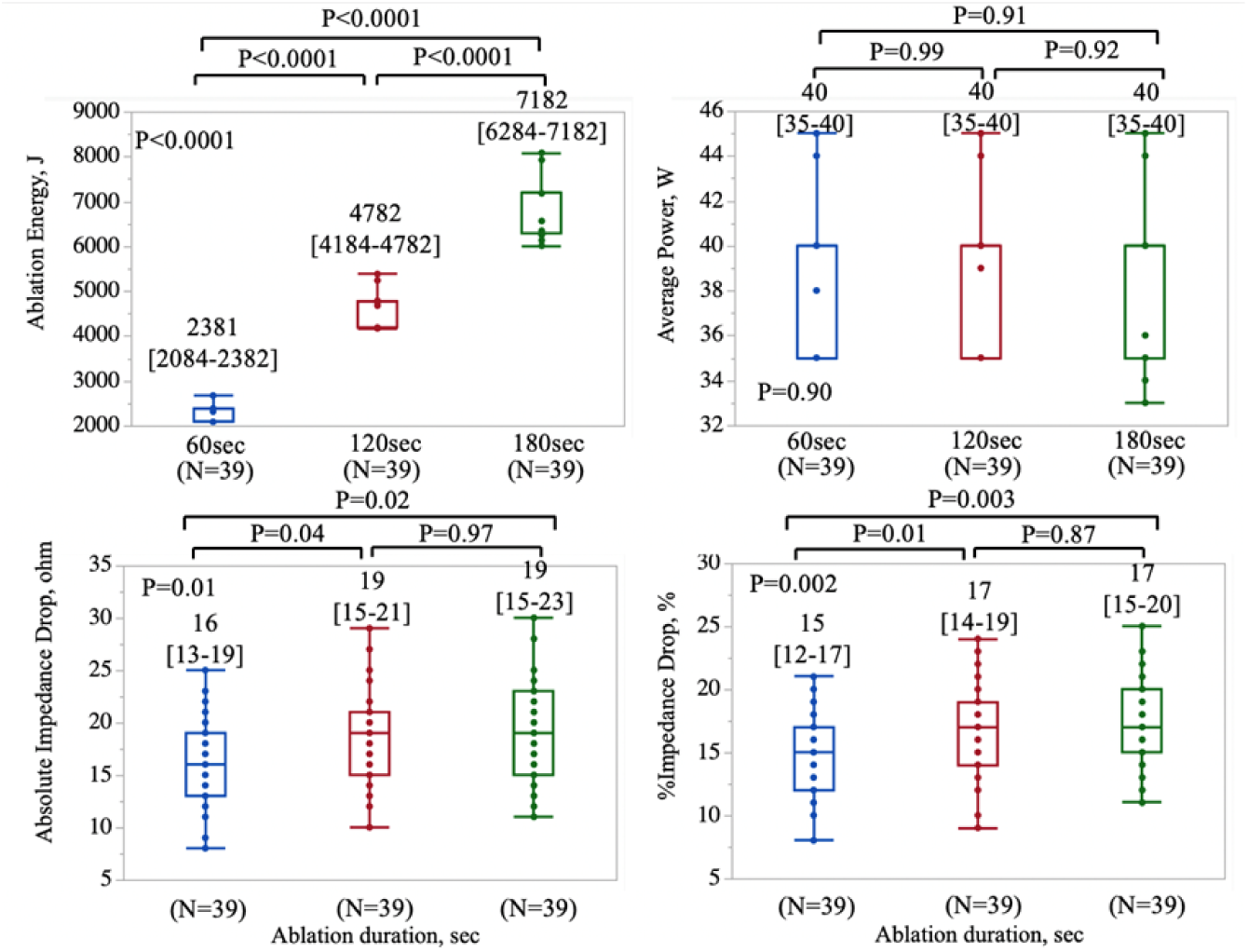
Comparison of lesion metrics and ablation parameters between three long RF-applications in ex-vivo model. **(A) Ablation duration vs Lesion metrics** **(B) Ablation duration vs Ablation parameters**

**Table 1.**

A. Ablation duration vs lesion metrics including all lesions
| Lesion metrics | 60s NS (n=48) | 120s NS (n=48) | 180s NS (n=48) | 60s HNS (n=48) | 120s HNS (n=48) | 180s HNS (n=48) |
| --- | --- | --- | --- | --- | --- | --- |
| Length, mm | 11.3[10.1-12.1] | 11.4[10.4-12.4] | 13.7[12.2-15] | 13.5[12.7-14.5] | 14.6[13.3-15.4] | 14.9[13.7-15.9] |
| Depth, mm | 6.0[5.2-6.6] | 5.6[5.2-6.3] | 7.7[7.0-8.4] | 7.5[6.7-8.3] | 8.5[8.1-9.2] | 8.6[7.6-9.5] |
| Volume, mm<sup>3</sup> | 404[331-529] | 410[319-506] | 819[572-1076] | 782[584-971] | 992[825-1137] | 1046[787-1328] |
| Area, mm<sup>2</sup> | 45.9[40.2-51.7] | 45.5[35.7-51.0] | 53.2[46.4-61.6] | 54.7[44.5-64.0] | 59.5[51.9-65.9] | 60.4[50.1-73.6] |
| Ablation parameters |  |  |  |  |  |  |
| Energy, J | 2382[2084-2679] | 2382[2084-2679] | 4306[4184-4782] | 4782[4184-5198] | 6284[5137-6391] | 6785[6284-7182] |
| Average power, W | 40[35-45] | 40[35-45] | 40[35-45] | 40[35-45] | 40[35-44] | 40[35-45] |
| Duration, sec | 60[60-60] | 60[60-60] | 120[120-120] | 120[120-120] | 125[180-180] | 180[180-180] |
| Max temp, °C | 34[31-35.8] | 34[32.3-36.8] | 35[32-38] | 35[32-37.8] | 35.5[34-39] | 35[32-38] |
| Absolute impedance drop, ohm | 20[16-23] | 16[13-19] | 22.5[19.3-28] | 19[15.3-21.8] | 24.5[20-28.8] | 19[16-23] |
| %Impedance drop, % | 16.5[14.0-18.8] | 14.5[12-17] | 18.5[16.3-22] | 17[14-19.8] | 20[17-23] | 18[15-21] |
| Average CF, g | 14.5[10-20] | 16[10-20] | 14.5[10-20] | 15.5[10-20] | 14.5[10-19] | 15.5[10-20] |

B. Ablation duration vs lesion metrics excluding steam-pop lesions
| Lesion metrics | 60s NS (n=48) | 120s NS (n=44) | 180s NS (n=39) | 60s HNS (n=46) | 120s HNS (n=37) | 180s HNS (n=27) |
| --- | --- | --- | --- | --- | --- | --- |
| Length, mm | 11.4[10.4-12.4] | 13.5[12.4-14.5] | 14.8[13.6-15.9] | 11.1[10.1-12.0] | 13.3[12.1-14.6] | 14.9[13.5-15.4] |
| Depth, mm | 5.6[5.2-6.3] | 7.5[6.7-8.2] | 8.6[7.6-8.9] | 6.0[5.2-6.4] | 7.3[7.0-8.2] | 8.5[8.1-8.9] |
| Volume, mm<sup>3</sup> | 410[319-506] | 775[580-970] | 1005[778-1214] | 397[324-481] | 682[559-911] | 1025[831-1109] |
| Area, mm<sup>2</sup> | 45.5[35.7-51.0] | 54.1[44.5-64.0] | 60.1[49.2-70.8] | 45.7[40.0-51.4] | 51.3[44.5-60.7] | 58.3[51.8-65.8] |
| Ablation parameters |  |  |  |  |  |  |
| Energy, J | 2382[2084-2679] | 4782[4184-5343] | 7182[6284-7182] | 2382[2084-2679] | 4774[4184-5081] | 6285[6284-7182] |
| Average power, W | 40[35-45] | 40[35-44.8] | 40[35-40] | 40[35-45] | 40[35-42.5] | 35[35-40] |
| Duration, sec | 60[60-60] | 120[120-120] | 180[180-180] | 60[60-60] | 120[120-120] | 180[180-180] |
| Max temp, °C | 34[32.3-36.8] | 34.5[32-37] | 34[32-37] | 34[31-35.3] | 34[31.5-36] | 35[32-41] |
| Absolute impedance drop, ohm | 16[13-19] | 19[15-21] | 19[15-23] | 20[16-22.3] | 22[19.5-24.5] | 20[19-25] |
| %Impedance drop, % | 14.5[12-17] | 17[14-19] | 17[15-20] | 16.0[14.0-18.0] | 18[16.5-20.5] | 17[16-22] |
| Average CF, g | 16[10-20] | 15.5[10-20] | 11[10-20] | 14.5[10-20] | 11[10-20] | 10[10-19] |

C. Ablation power vs lesion metrics including all lesions
| Lesion metrics | 35W NS (n=48) | 40W NS (n=48) | 45W NS (n=48) | 35W HNS (n=48) | 40W HNS( n=48) | 45W HNS (n=48) |
| --- | --- | --- | --- | --- | --- | --- |
| Length, mm | 12.6[10.7-13.5] | 13.8[11.6-15.0] | 14.2[12.4-15.3] | 12.2[11.5-13.9] | 13.4[11.2-15.2] | 13.6[12.2-15] |
| Depth, mm | 6.8[5.3-7.6] | 7.5[6.2-8.4] | 8.0[6.4-8.9] | 7.2[6.1-8.4] | 7.7[6.6-8.5] | 8.0[6.5-9.1] |
| Volume, mm<sup>3</sup> | 562[341-749] | 769[464-1034] | 890[554-1160] | 580[449-864] | 770[463-1091] | 806[540-1125] |
| Area. mm<sup>2</sup> | 46.9[36.4-56.1] | 50.5[43.7-62.6] | 59.4[50.7-72.7] | 47.8[41.9-57.0] | 53.6[47.3-62.7] | 53.4[48.4-66.9] |
| Ablation parameters |  |  |  |  |  |  |
| Energy, J | 4184[2084-6284] | 4782[2382-7181] | 5380[2680-6371] | 4184[2084-6248] | 4748[2382-5148] | 3387[2679-5380] |
| Average power, W | 35[35-35] | 40[40-40] | 45[45-45] | 35[35-35] | 40[40-40] | 45[45-45] |
| Duration, sec | 120[60-180] | 120[60-180] | 120[60-153] | 120[60-180] | 120[60-125] | 75.5[60-120] |
| Max temp, °C | 34[32.3-35] | 34[32-37] | 35[32-38] | 34[32-35.8] | 35[32-39] | 35[32.3-37.8] |
| Absolute impedance drop, ohm | 17[13.5-20.8] | 19[15-20] | 18.5[15-23.8] | 20[17.3-23] | 21[17-29.5] | 24[21-29] |
| %Impedance drop, % | 16[12-18] | 17[14-18] | 17[14-21] | 17[15-18.8] | 18[15-23] | 20[18-23.8] |
| Average CF, g | 16[10-20] | 15.5[10-20] | 15.5[10-20] | 14.5[10-20] | 14.5[10-19] | 14.5[10-19] |

D. Ablation power vs lesion metrics excluding steam-pop lesions
| Lesion metrics | 35W NS (n=48) | 40W NS (n=45) | 45W NS (n=38) | 35W HNS (n=46) | 40W HNS (n=37) | 45W HNS (n=27) |
| --- | --- | --- | --- | --- | --- | --- |
| Length, mm | 12.6[10.7-13.5] | 13.7[11.5-15.0] | 13.4[12.3-15.0] | 12.2[11.5-13.9] | 12.8[10.9-15.0] | 13.1[11.2-14.7] |
| Depth, mm | 6.8[5.3-7.6] | 7.4[6.2-8.3] | 7.5[6.2-8.6] | 7.1[6.1-8.2] | 7.1[6.2-8.3] | 7.2[6.0-8.5] |
| Volume, mm<sup>3</sup> | 562[341-749] | 774[463-1029] | 782[506-1066] | 571[448-864] | 668[410-1072] | 650[419-946] |
| Area. mm<sup>2</sup> | 46.9[36.4-56.1] | 49.7[43.8-63.0] | 56.4[49.3-71.8] | 47.0[41.5-55.9] | 52.0[45.8-62.6] | 50.9[45.5-63.1] |
| Ablation parameters |  |  |  |  |  |  |
| Energy, J | 4184[2084-6284] | 4782[2382-7182] | 5380[2680-5621] | 4184[2084-6284] | 4774[2382-5616] | 2680[2679-5380] |
| Average power, W | 35[35-35] | 40[40-40] | 45[45-45] | 35[35-35] | 40[40-40] | 45[45-45] |
| Duration, sec | 120[60-180] | 120[60-180] | 120[60-135] | 120[60-180] | 120[60-150] | 60[60-120] |
| Max temp, °C | 34[32.3-35] | 34[32-36] | 34[32-37.3] | 34[32-36] | 35[31-39.5] | 34[30-36] |
| Absolute impedance drop, ohm | 17[13.5-20.8] | 19[15-20] | 17[14.8-22.3] | 20[17-23] | 20[16-20] | 22[20-24] |
| %Impedance drop, % | 16[12-18] | 17[14-18] | 16.5[13.8-20] | 17[15-18.3] | 17[14-21] | 18[17-20] |
| Average CF, g | 16[10-20] | 12[10-20] | 15.5[10-20] | 14.5[10-20] | 11[10-19.5] | 10[10-20] |

### Confirmation in In-vivo experiment

Although Swine 1,3, and 4 have completed the entire RF-application protocol, 35W/180s, 40W/120s, 40W180s with HNS, and 45W/180s with NS were not performed in Swine 2 because the experiment was terminated due to animal death. As shown in Figure 5, six steam-pops were only observed in RF-applications irrigated with HNS (0/35, 0% in NS vs 6/21, 28.6% in HNS, P<0.0001) even though aggressive settings (35W/180s, 40W/120s. 40W/180s with HNS) were not performed in the Swine 2 because this animal died before the complete protocol was achieved. Steam-pops were observed in the setting where the catheter was forced to be parallelly placed at the tissue with a relatively larger CF due to a small ventricular chamber, and steam-pops were non-audible in half of them.

**Figure 5.**
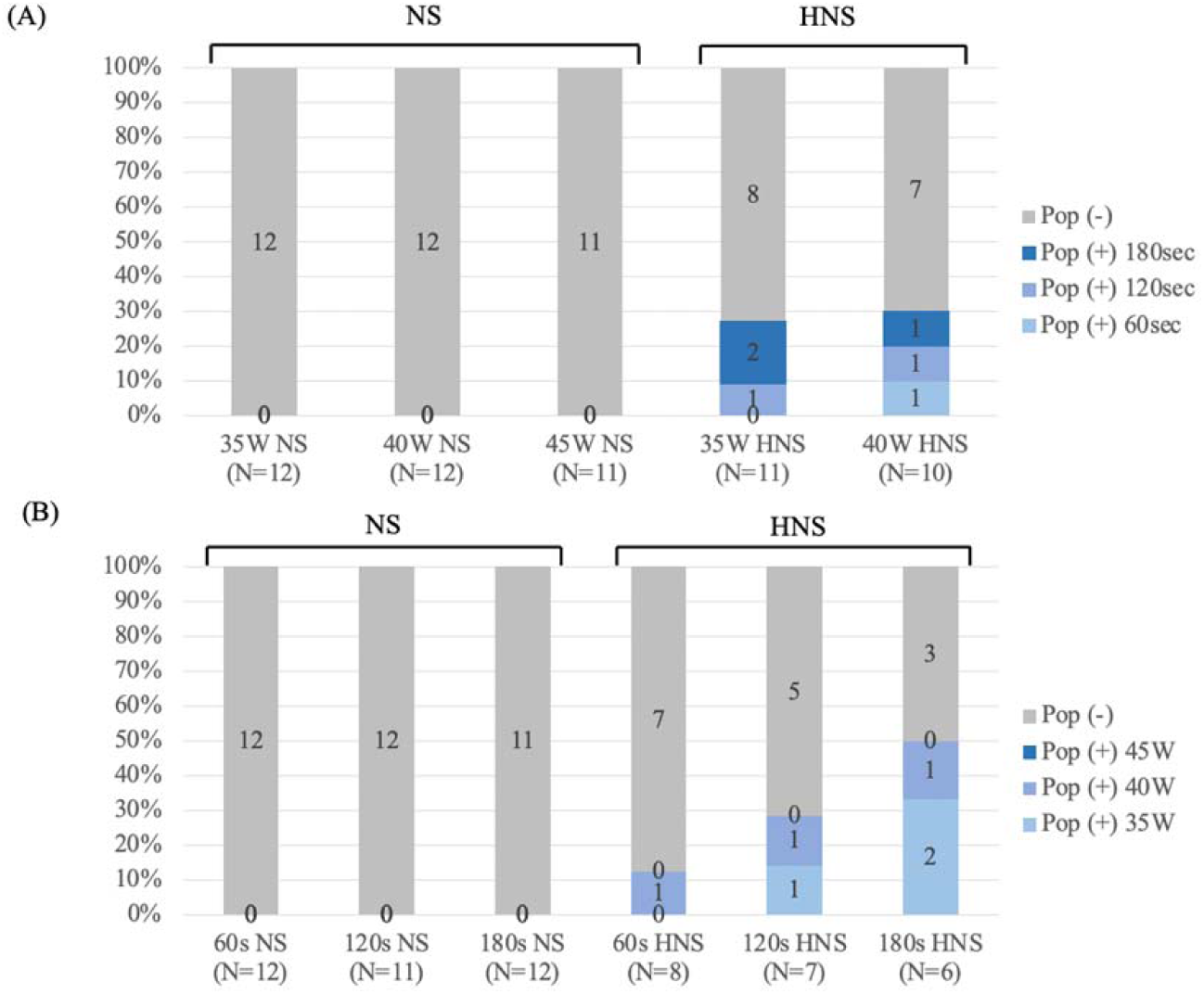
Incidence of steam-pops in in-vivo condition. **A. Ablation power vs steam-pops** **B. Ablation duration vs steam-pops**

To avoid the bias of the comparison of lesion metrics between the NS group and the HNS group, ablation settings causing steam-pops and those not being performed because of the animal death in one cohort were also excluded from the other cohort. In total, 17 lesions in the NS-group and those in the HNS-groups were compared, showing that the length, depth, volume, and surface area of the lesion did not differ between the two groups as shown in Figure 6A and 6B, which were similar to the findings from the ex-vivo study. Power titration due to the temperature guard function was more frequently observed in HNS-irrigation (10/17, 59%) than in NS-irrigation (5/17, 29%) (Supplemental figure 1), probably leading to lower average energy in the HNS-group (Figure 6C).

**Figure 6.**
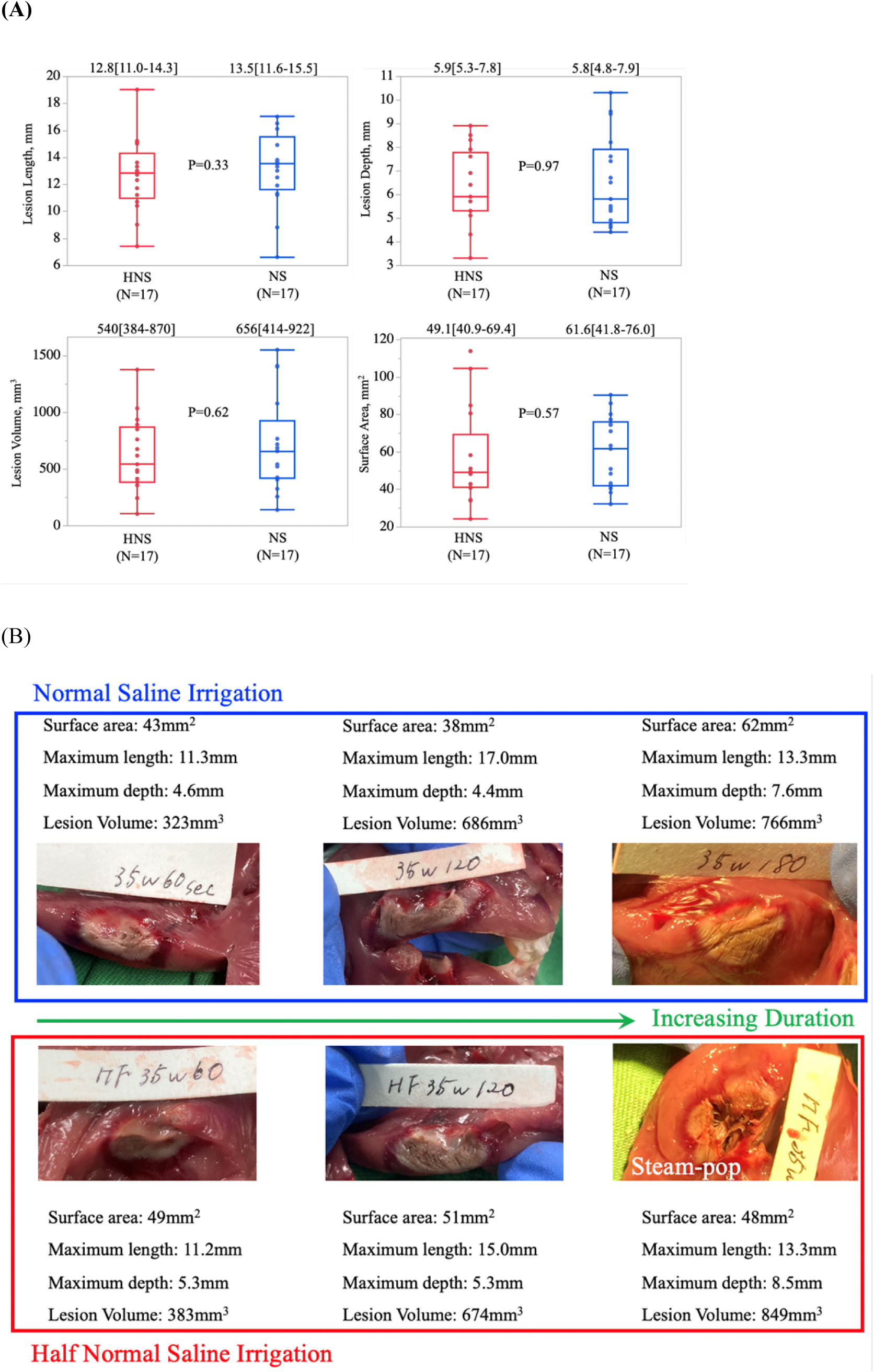
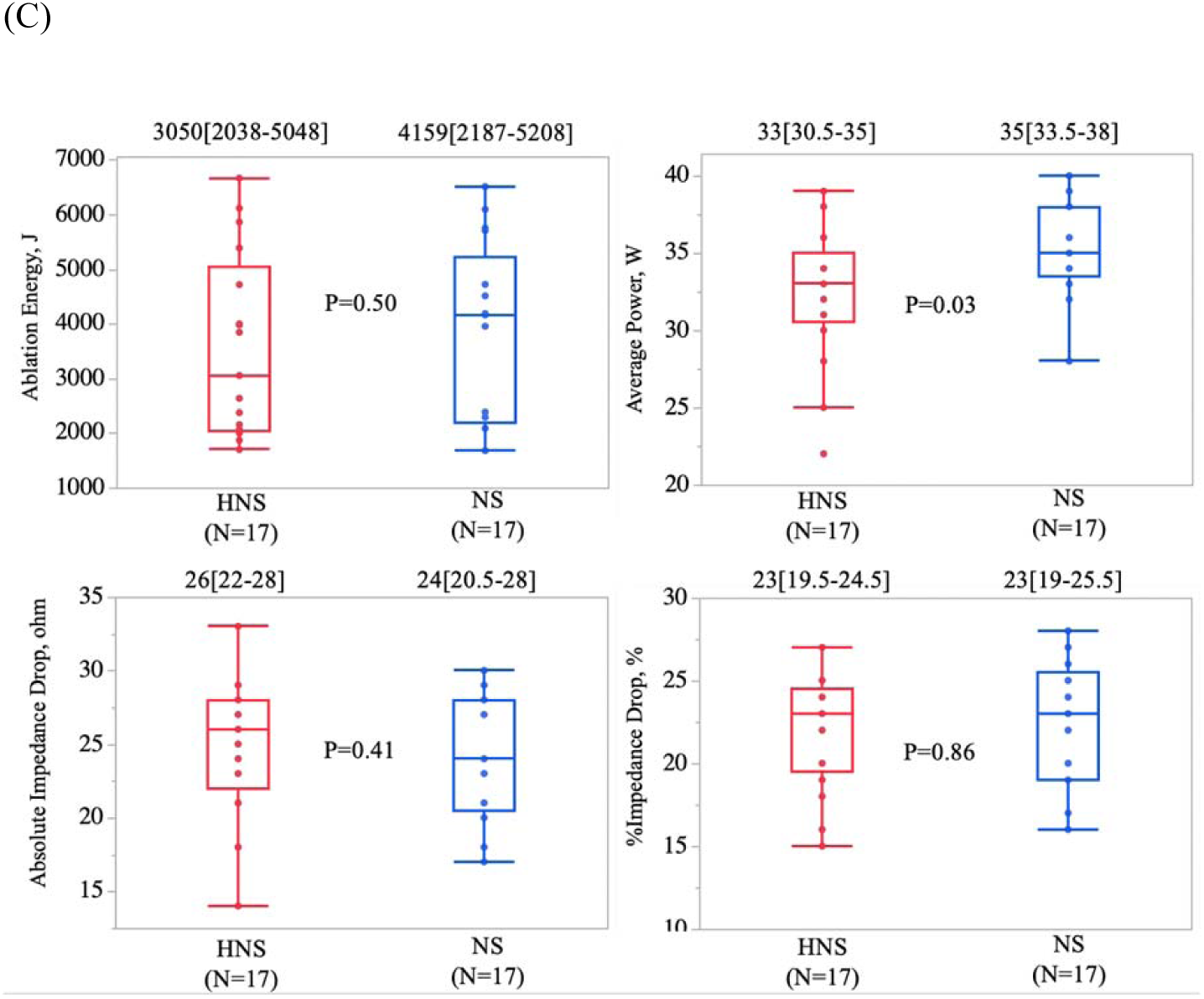
Comparison of lesion metrics and ablation parameters between HNS-irrigation and NS-irrigation in ex-vivo model. **(A) HNS-irrigation vs NS-irrigation: Lesion metrics** **(B) HNS-irrigation vs NS-irrigation: demonstration of lesion metrics in different ablation settings** **(C) HNS-irrigation vs NS-irrigation: Ablation parameters**

The impact of increasing ablation duration with this catheter was demonstrated in the Figure 7. Despite the absence of one application of 45W/180s due to the animal death and the small number of total lesions, a remarkable tendency of increasing lesion size was observed as the ablation duration increased (Figure 7A and B). Although impedance-drop may be larger in 120s-group compared to 60s-group, no difference was observed between the 120s group and 180s group (Absolute impedance drop; 25[23-28]ohm in 120s group vs 24[23-28]ohm in 180s group, P=0.87, and %impedance drop; 24.5[21.3-25.8]% in 120s group vs 25[24-26]% in 180s group P=0.50).

**Figure 7.**
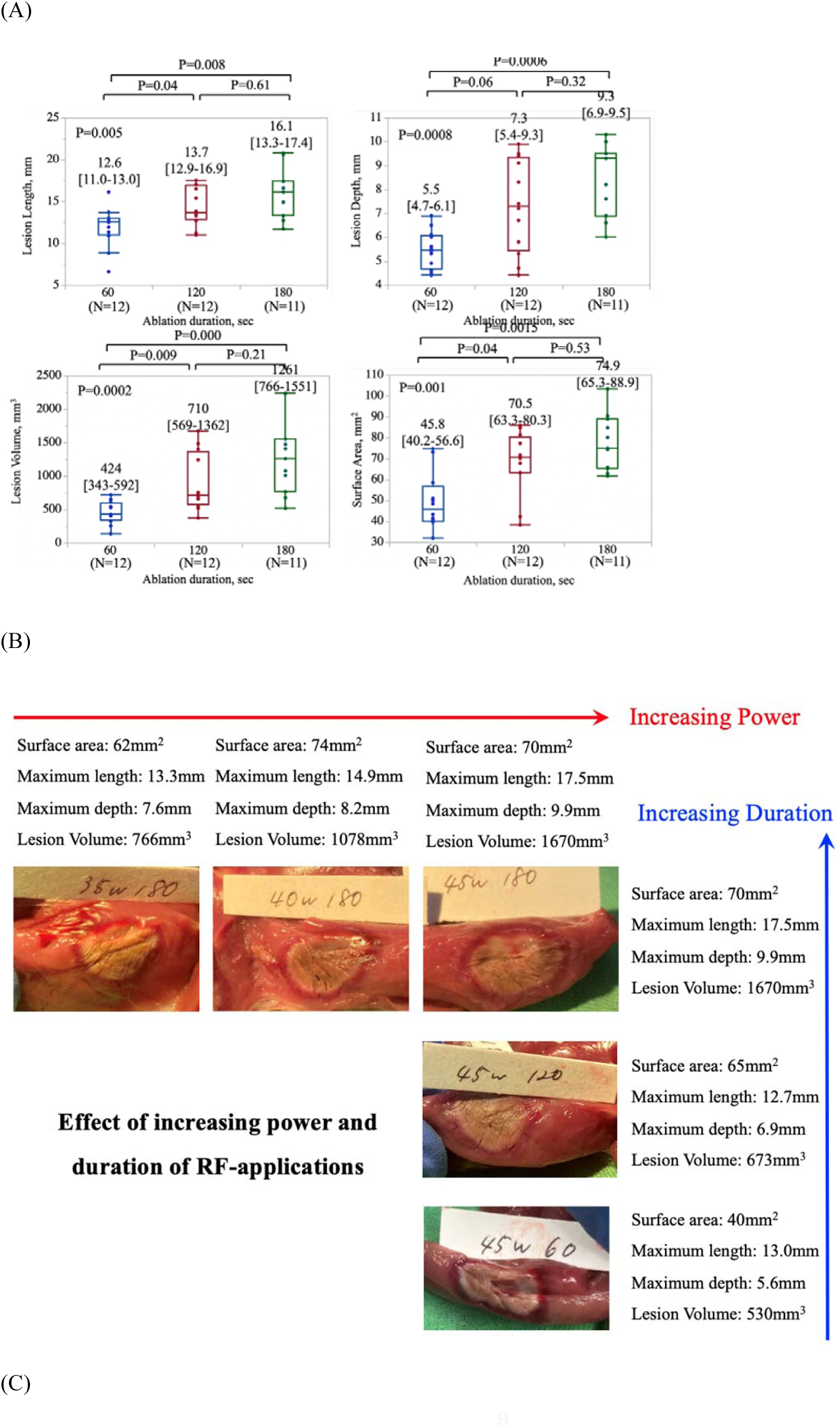
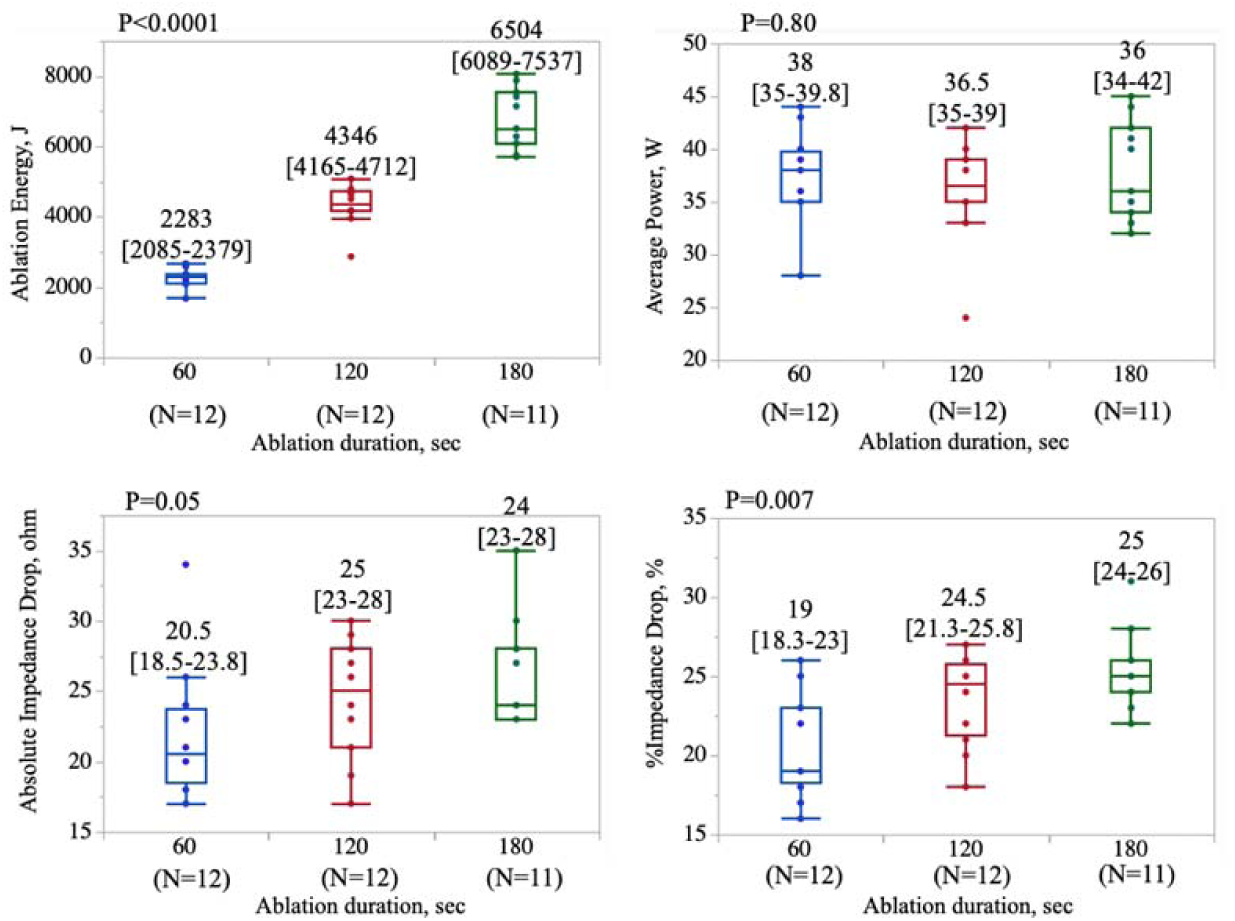
Comparison of lesion metrics and ablation parameters between three long RF-applications in in-vivo model. **(A) Ablation duration vs Lesion metrics** **(B) Demonstration of different lesion metrics in different ablation settings** **(C) Ablation duration vs Ablation parameters**

### Impedance variation vs steam-pops

In the ex-vivo experiment, out of 288 lesions, 47 (16.4%) steam-pops were observed including 13 with HNS-irrigation and 34 with NS-irrigation. By ROC curve analysis using the ex-vivo data including both NS-irrigation and HNS-irrigation, the area under the curve (AUC) of the %Impedance-drop in predicting steam pops was 0.861 (Figure 8A). The optimal cut-off value of %Impedance-drop was 20% with the sensitivity, specificity, positive predictive value (PPV), and negative predictive value (NPV) of 78.3%, 80.6%, 43.4%, and 95.1%, respectively. When the analysis was performed only in the NS-irrigation group, the AUC and the optimal cut-off value of %Impedance-drop were 0.856 and 22% with the sensitivity, specificity, PPV, and NPV of 61.5%, 93.1%, 47.1%, and 96.1%, respectively (Figure 8B).

**Figure 8.**
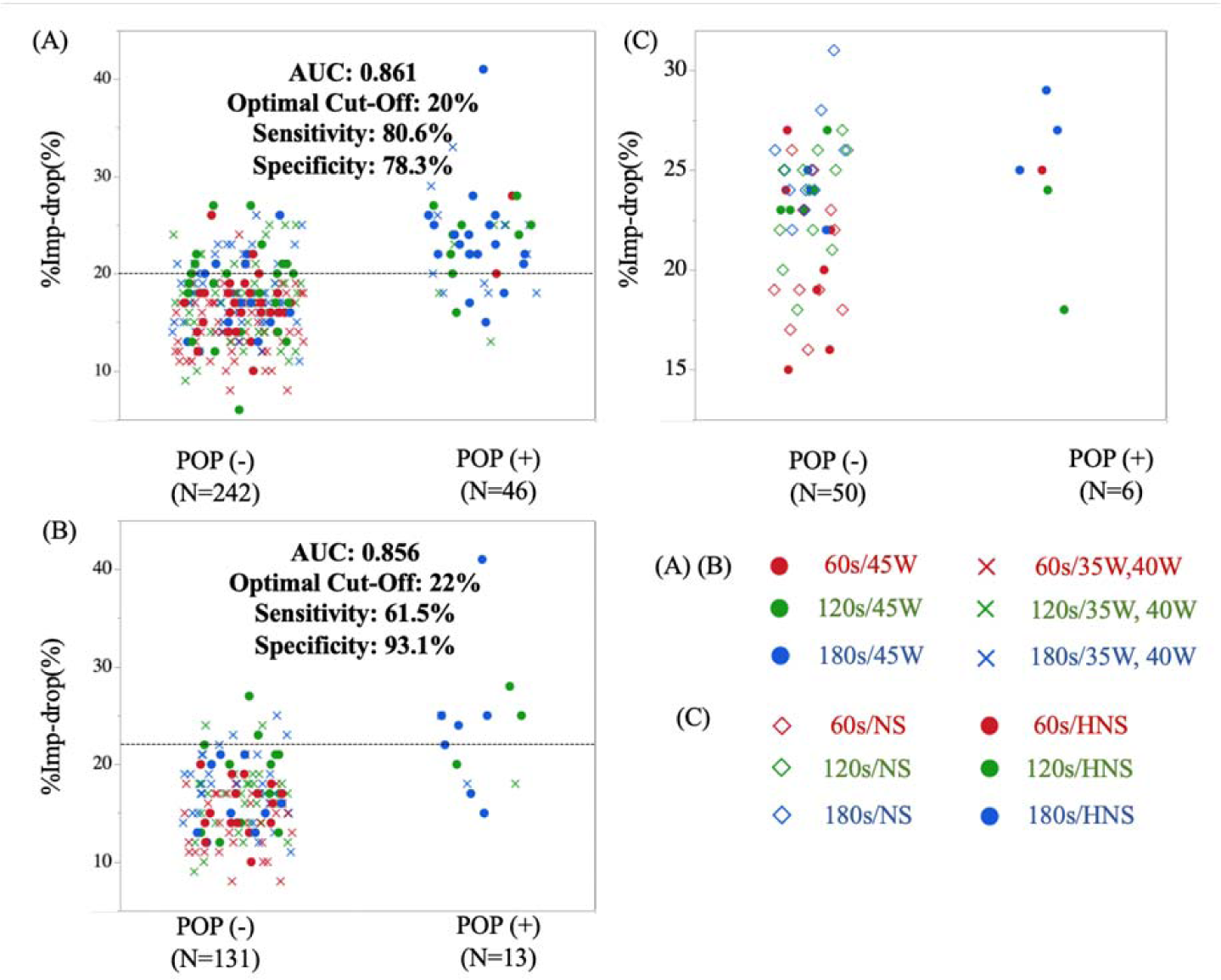
Accuracy and cut-off values of impedance drop for the prediction of steam-pops. (A) Optimal cut-off values to predict steam-pops in total ablation lesions (ex-vivo) (B) Optimal cut-off values to predict steam-pops in lesions with NS-irrigation (ex-vivo) (C) Optimal cut-off values to predict steam-pops in total ablation lesions (in-vivo)

Figure 8C showed the comparison of impedance variation during RF-applications between lesions with steam-pops (n=6) and without (n=50), including both NS-irrigation and HNS-irrigation. The AUC and the optimal cut-off value of %Impedance-drop was 0.677 and 24% with the sensitivity, specificity, PPV, and NPV of 83.0%, 52.0%, 17.2%, and 96.3%, respectively. However, all 6 steam-pops were observed with HNS-irrigation, and no steam-pops were observed even with 45W/180s RF-applications in the NS-irrigation group. suggesting that the cut-off value of %Impedance-drop=20%, being derived from the ex-vivo analysis, may be still used as an appropriate cut-off with a sufficient safety margin in in-vivo setting using NS-irrigation.

## Discussion

We investigated the strategy to enlarge lesion size in temperature control mode using an CF-sensing irrigation catheter with a flexible laser-cut tip with unique irrigation in producing larger lesions safely and efficiently. We demonstrated that:

1. Lesion size proportionally increased as the RF duration increases in power settings of 35, 40, and 45W. The risk of steam-pops may also increase as the duration and power increases in the ex-vivo study. Although the in-vivo experiment showed the same tendency, the risk of steam-pops may be limited at least in NS-irrigation.
2. HNS-irrigation showed significantly larger impedance-drop and increased the risk of steam-pops, but did not generally increase the lesion size, which were demonstrated in both ex-vivo and in-vivo studies.
3. The optimal cut-off value of %Impedance-drop=20% may be used as a reliable marker to predict the incidence of steam-pops with a high NPV even in a long RF-applications.

### How to create deep lesions

As far as we know, this is the first study to discuss some potential methods to increase the lesion size without increasing a risk of steam-pops using a catheter with a novel laser-cut flexible tip.

It has been challenging to eliminate the arrhythmogenic regions contained deep within myocardium^2^, such as interventricular septum, left ventricular summit, non-septal midmyocardial sites, and papillary muscles^3^.

Although longer RF applications have been believed to enlarge the lesion size, standard irrigation catheters are generally power-controlled, and systematic experiments to answer this question has not been performed so far. In the present study, we clearly demonstrated that longer RF applications (60s, 120sec, and 180sec) might proportionally increase the lesion size in moderate-to high-power settings (35W, 40W, and 45W) in temperature control mode using the TactiFlex^TM^ catheter.

In contrast, HNS-irrigation may not safely achieve the deeper lesions, although it has been believed to increase the lesion size by increasing the current density into the tissue in power-control ablation. Although the impedance drop was larger in the HNS-irrigation group, the lesion volume was not always enlarged, and the steam-pops significantly increased. Theoretically, irrigation with a highly resistive substance may serve as an “insulator”, preventing the radiofrequency energy from dispersing into the surrounding blood pool, and instead, current density under the tissue more rapidly increases. This acute reaction may be associated with a more rapid and larger impedance drop. Despite the larger current flowing into the tissue, the lesion size may not always statistically increase, probably due to the power titration attributed to the rapid tissue temperature increase or steam-pops before the activation of the power titration. At least in in-vivo settings or clinical settings, RF-applications up to 180s with the power of 35-45W may increase the lesion size with a limited risk of steam-pops.

### Impedance variation for a prediction of lesion size and steam-pops

The proportional effect of the longer RF-application duration on the lesion volume did not reach significance but at least showed the tendency in in-vivo experiment. Further, this effect was remarkably demonstrated in the ex-vivo experiment where a sufficient number of lesions for statistical significance were achieved.

However, interestingly, the impedance variation such as the absolute impedance drop and %impedance drop did not proportionally increase, especially as there was no difference in impedance variation between 120s applications and 180s applications in both ex-vivo and in-vivo experiments. It may be concluded that the impedance variation might not be used for the predictor of the lesion size in long duration RF-applications. For the prediction of the steam-pops, the cut-off %impedance drop>20% highly predicted the risk of steam-pops with high NPV=95.1% in RF-applications with both NS-and HNS-irrigations. When we focus on RF-applications with NS-irrigation, the cut-off %impedance drop>22% also highly predicted the risk of steam-pops with high NPV=96.1%. We previously reported that the cut-off value of 20% was optimal to predict steam-pops in ex-vivo experiment in RF-applications up to 60s. Further, the clinical data showed that this value could be used with a sufficient safety margin, but mainly proved by the RF-applications with NS-irrigation. Although we believe that %impedance-drop=20% is still a reasonable cut-off value to safely predict steam-pops with a high NPV even with the longer RF-applications >60s, some applications in the ex-vivo experiment and those with HNS-irrigation even in in-vivo experiments produced steam-pops even with the lower %impedance-drop. It should be stressed that in the condition with parallel catheter placement with high-contact, HNS-irrigation, or high initial impedance when the large part of the catheter tip is considered to be covered with cardiac tissue such as trabeculation, RF-applications should be carefully performed because the large part of the electric current may rapidly flow into the tissue without dissipating to the blood-pool, resulting in a steam-pops before reaching the %impedance=20%.

### Clinical implications

To achieve larger and deeper lesions with this catheter, extending the duration of RF application may be recommended rather than using HNS for irrigation. Lesions become larger and no steam-pops were observed even with 45W/180s applications in the in-vivo experimental model. However, this maximum setting should be carefully performed because some steam-pops were observed in ex-vivo setting, although the ex-vivo environment may tend to show more frequent steam-pops due to the less convective cooling effect.

### Limitations

These experiments were performed in an ex-vivo model or an in-vivo model but with healthy swine ventricular myocardium, and therefore the results may not necessarily translate to clinical human pathology. However, a human study with histopathology may not be feasible. Although variations in optimal catheter orientation in the in-vivo model may have affected lesion sizes, this is a common clinical problem in reality, and one which we attempted to control as much as was feasible in a beating heart model.

## Conclusions

In temperature control mode, longer RF applications may be recommended instead of using half-normal saline irrigation to safely achieve deeper lesions.

## Data Availability

DATA AVAILABILITY: The data underlying this article will be shared on reasonable request to the corresponding author.

## Acknowledgement

We appreciate a technical support for this experiment from Mr. Masashi Uemoto, Mr. Takenori Yamada, Mr. Kitabata, Mr. Ryoto Shiga, and Miss. Lisa Sakurai from Abbott Medical Japan LIS.

## Supplemental figure

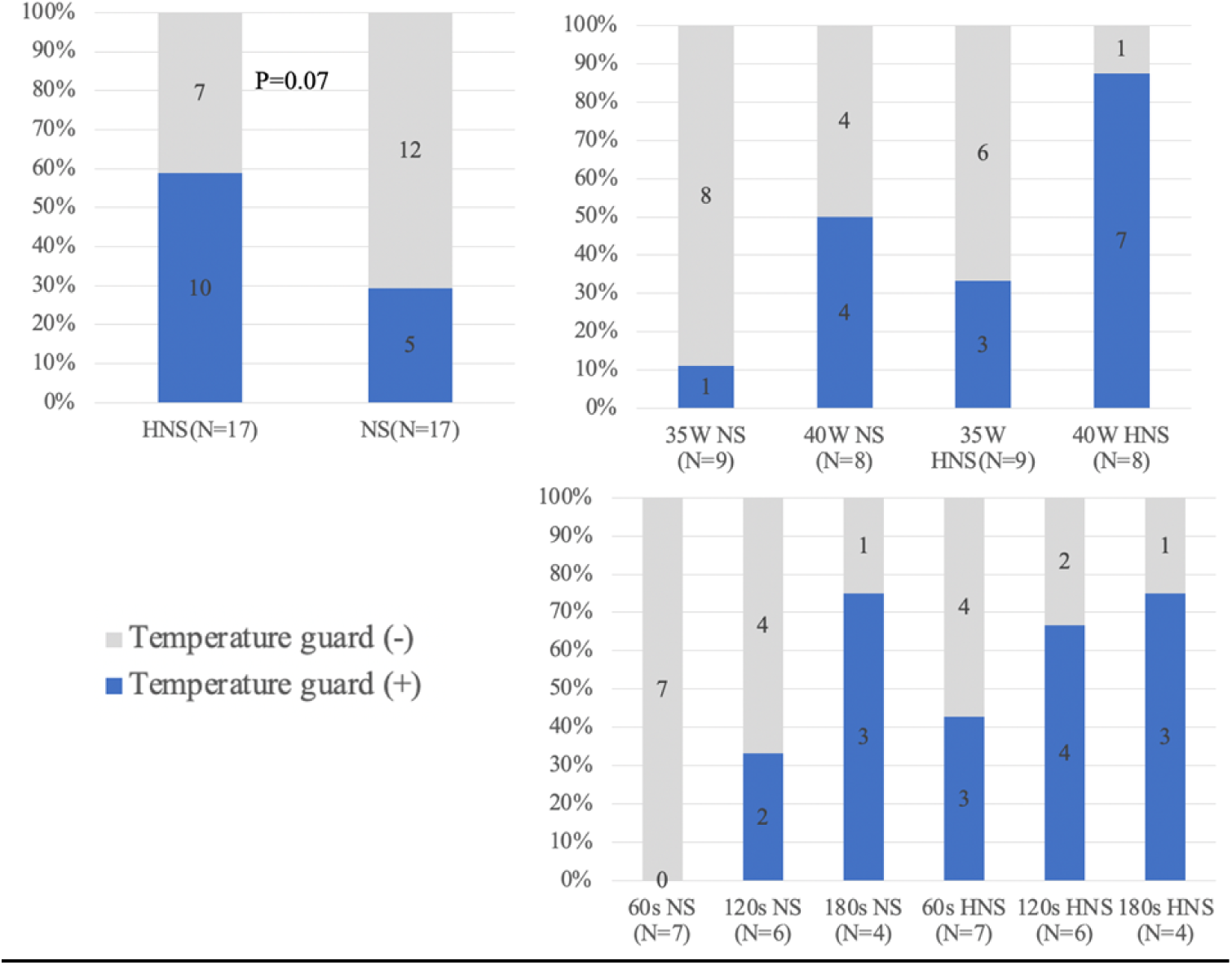

